# A deep learning approach for metabolic rate prediction

**DOI:** 10.1101/2022.10.24.22281456

**Authors:** Ioannis Kontopoulos, Christina-Ariadni Valagkouti, Panagiotis Kontopoulos

**Affiliations:** Department of Informatics and Telematics, Harokopio University of Athens, Greece; Space Medicine and Life Sciences, SGA Council, Austria; Department of Informatics and Telecommunications, National and Kapodistrian University of Athens, Greece

**Keywords:** deep learning, neural networks, metabolic rate prediction, oxygen consumption rate, heart rate

## Abstract

The sudden increase of wearable devices has led to the generation of an abundance of data. As a result, researchers can use such data to perform analyses and generate recommendations. A crucial factor in the research field of nutrition and dietetics is the accurate measurement of metabolic rate, as it can be used to estimate several other variables, e.g. calorie expenditure. Nonetheless, limited studies have been conducted to examine the use of machine learning models for metabolic rate prediction based on data generated from wearable devices. Therefore, in this paper, a neural network architecture is proposed, able to predict a subject’s metabolic rate exploiting only such data. Experimental results demonstrated that the proposed methodology can outperform conventional algorithms in prediction accuracy of real-world data. Furthermore, results indicated that the trivial time taken for the network to predict the metabolic rate makes it suitable for wearable devices deployment.

## 1 Introduction

Data on human energy expenditure rate is of interest to a range of scientific fields. Ergonomics studies rely on energy expenditure measurements in order to determine the workers’ strain levels, obesity studies use them to investigate the human body’s behaviour and responses, and nutrition counseling professionals involve them in their work routine. Golden standard methods include measurement of oxygen consumption rate (*V O*_2_), the doubly labelled water method, and direct calorimetry. While providing a high level of accuracy, these methods for measuring metabolic rate include cumbersome paraphernalia and sometimes elevated prices, that are obstructing their widespread application.

The measurement of heart rate (HR) is a relatively simple process that can be carried out, on varying levels of accuracy, by non-invasive, cost-effective wearable devices. Moreover, the relationship between *V O*_2_ and HR during exercise has been observed to be linear Bot and Hollander [2000], Schantz et al. [2019] and the HR measurement method is even recommended as a method to assess the energy costs of jobs or activities within ISO 8996:2004^1^. However, the same ISO standard states that the relationship is linear only within a specific range, and under defined conditions. Specifically, the method of determining the metabolic rate by “heart rate measurement under defined conditions” is estimated to have a *±*10% accuracy and to carry a medium error risk. It is also highlighted that, according to ISO 8996:2004, one of the main factors that influence the method’s accuracy is “the accuracy of the relationship between heart rate and oxygen uptake, as other stress factors also influence the heart rate”.

The linear relationship between HR and *V O*_2_ and the simplicity of HR measurement acquisition are being used by sport watch companies, which typically develop a proprietary, traditional algorithm that takes into account anthropometric variables, maximal HR (*HR*_*max*_) and maximal *V O*_2_ (*V O*_2*max*_) for calculating energy expenditure, through an equation Crouter et al. [2004]. However, such an approach ignores the limitations stated in ISO 8996:2004, resulting to some degree of inaccuracy, especially when it comes to high intensity exercise Cooper and Shafer [2019], Crouter et al. [2004], Erdogan et al. [2010], Roos et al. [2017]. Thus, in contrast to a traditional algorithm, we propose the use of Deep Learning (DL), which would be trained on sets of anthropometric, HR, and *V O*_2_ measurements, and would subsequently be able to produce a *V O*_2_ estimation by working with new input. To the best of our knowledge, limited studies have been conducted demonstrating that a DL approach is capable of estimating *V O*_2_ with greater accuracy than a traditional algorithm.

The main contributions of our research are the following:

- Neural Networks (NNs) are exploited to predict *V O*_2_ at any given time point. NNs have demonstrated an increased accuracy in several research domains.
- The subject’s *HR* and speed when running are used to predict *V O*_2_. These two variables can be easily obtained by commercial wearables, making the proposed approach deployable.
- The proposed approach is compared against other conventional techniques that are used to estimate *V O*_2_.

The rest of the paper is structured as follows. Section 2 presents the literature review in the respective field of research. Section 3 describes in detail the methodologies that will be evaluated. Specifically, Sections 3.1 and 3.2 describe the two methodologies that will be used as baselines and Section 3.3 describes the proposed *DL* approach. Finally, Section 4 presents the experimental evaluation and Section 5 concludes the merits of our work.

## 2 Related Work

Neural Networks (NNs) have been widely used over the years in numerous applications and research domains Abiodun et al. [2018], ranging from power electronics and motor drives Bose [2007] to geographic data and information Kontopoulos et al. [2021], and healthcare Makris et al. [2020]. With the passing of time, neural networks continue to find new applications to be deployed to. Consequently, NNs have found their way in the research field of nutrition and dietetics with several studies having been conducted Zecchin et al. [2012], D.Thangamani and P.Sudha [2014]. A crucial element in nutrition and dietetics is the metabolic rate, because it can be used to estimate several other variables such as calorie expenditure and thermal comfort. Metabolic rate is most commonly calculated indirectly via measurements of *V O*_2_. Therefore, it is important to develop a methodology able to accurately predict *V O*_2_ values.

In an attempt to predict the thermal comfort of subjects which is based on the metabolic rate, authors in Na et al. [2019], Na and Kim [2019] used a Kinect camera to obtain images and a wearable device to collect *HR* data. Then, they trained a *DL* model to predict the subject’s *HR* based on the collected images. The predicted *HR* was later used in combination with the subject’s weight, sex and age to estimate the metabolic rate. Similarly, Liu et al. Liu et al. [2021] attempted to assess the subject’s thermal comfort, by developing a vision-based approach to estimate individual clothing insulation rate and metabolic rate; the two critical factors to assess personal thermal comfort level. To this end, they implemented a Convolutional Neural Network (CNN) to recognize the subject’s clothes type and activity type (e.g., running) through a thermal camera and estimate the skin and the clothes temperature. Contrary to these studies, our proposed approach tries to directly predict the metabolic rate using NNs. In an another attempt to investigate the use of NNs, authors in Freedson et al. [2011] conducted a survey, evaluating algorithms able to predict metabolic equivalents (METs) and activity types. The methodologies in the survey used as input either accelerometer data and activity types in order to predict METs, or accelerometer data in order to predict activity types. The main difference between our proposed approach and the aforementioned ones is that our methodology tries to directly predict *V O*_2_ instead of predicting METs and activity types. The study that has the most similarities to the proposed approach was conducted by authors in Rothney et al. [2007], where they developed a feed-forward/back-propagation NN with one hidden layer (12*×* 20*×* 1 nodes). Their NN takes as input 10 signal features extracted from accelerometers placed at the hip of the subjects. Their proposed NN was able to predict the subject’s metabolic rate more accurately than other methodologies which monitored the subject’s acceleration. Despite the fact that they used a NN for metabolic rate prediction, subjects needed to wear accelerometers to acquire data suitable for training. The main advantage of our approach lies in the fact that the proposed neural network architecture takes as input variables that can be obtained by wearables and smartphones, making it easier to be deployed and used.

## 3 Methodology

In this section, we describe the baseline methodologies used to predict *V O*_2_ from HR measurements. Specifically, in Sections 3.1 and 3.2, tow baseline methodologies are described. In Section 3.3, a new methodology is proposed and described for the prediction of *V O*_2_ from HR measurements.

### 3.1 Linear correlation

Generally, HR measurements can be used for *V O*_2_ estimation in free-ranging animals, including humans, if HR and *V O*_2_ are calibrated against each other in a controlled environment Butler et al. [2004]. The majority of sports watch developers treat the algorithms they use with confidentiality, however the history of the HR measurement method suggests that calibration occurs by generation of an HR-*V O*_2_ diagram Lundgren [1947]. The two variables are connected by a significant, linear relationship. A mixed model analysis has identified sex, weight, age, and fitness as factors that determine the slope of the HR-*V O*_2_ relationship Keytel et al. [2005], implying a unique, one-to-one linear HR-*V O*_2_ relationship for every individual.

In this research, we used a one-to-one linear correlation between *HR* and *V O*_2_ as a baseline. To calculate the *V O*_2_ at the moment of interest (*V O*_2*t*_), we need to take into account the subject’s *V O*_2*max*_, its *HR*_*max*_, and its HR at the moment of interest (*HR*_*t*_). Based on the one-to-one correlation between HR and *V O*_2_, we can infer that the *V O*_2*t*_ is a percentage of *V O*_2*max*_, which can be extrapolated by the value that represents the percentage of *HR*_*max*_ that is *HR*_*t*_:

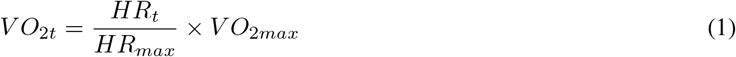

### 3.2 A Theoretical Method of Using Heart Rate to Estimate Oxygen Uptake During Exercise

In an effort to determine the slope more accurately, Pettitt and his team Pettitt et al. [2007] presented a methodological paper describing a step-by-step procedure to estimate *V O*_2_ at a target *HR*. The method includes a *V O*_2_ reserve coefficient (*RSV*_*c*_), which reflects the increase in *V O*_2_ for every *HR* beat above the *HR* at rest (*HR*_*rest*_). *RSV*_*c*_ is obtained by dividing the difference between *V O*_2*max*_ and *V O*_2_ at rest (*V O*_2*rest*_) by the difference between *HR*_*max*_ and *HR*_*rest*_. The resulting formula takes this form:

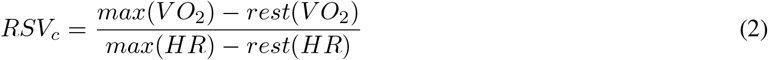

Subsequently, *RSV*_*c*_ can be used to determine *V O*_2_, in Pettitt’s case, at a target *HR*, and in our case, at *HR*_*t*_. The number of *HR* beats above *HR*_*rest*_, multiplied by *RSV*_*c*_, gives an estimation of the increase in *V O*_2_ that is a direct consequence of the increase in *HR*. Adding to this the *V O*_2*rest*_, the calculation for *V O*_2*t*_ is complete. The resulting equation is of this form:

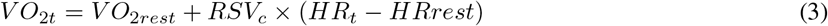

### 3.3 Neural Network

We propose a deep NN architecture that is able to accurately estimate the *V O*_2_, given the *HR*, and the speed at which the subject moves when running. Both these variables can be easily obtained by widely used, commercial smartwatches and smartbands, thus any algorithm that exploits these variables can be easily deployed. Therefore, the proposed NN architecture takes as input the subject’s current heart rate and speed and outputs the predicted oxygen uptake *V O*_2_. Table 1 illustrates the architecture used for the *V O*_2_ prediction. The proposed NN has 4 fully connected dense layers each one followed by a dropout of 0.4, meaning that 40% of the nodes of each layer will be randomly dropped. Furthermore, each layer uses the ReLu activation function due its reduced likelihood of vanishing gradient and its efficient computation. Finally, the last layer which outputs a single value that corresponds to the predicted oxygen uptake uses the sigmoid activation function. The Adam optimizer was used for our proposed NN architecture.

**Table 1:** The neural network architecture.

| Number of parameters: 11,153 |  |  |
| --- | --- | --- |
| Layer type | Number of nodes | # params |
| Dense | 16 | 48 |
| Dropout (0.4) | - | 0 |
| Dense | 32 | 544 |
| Dropout (0.4) | - | 0 |
| Dense | 64 | 2112 |
| Dropout (0.4) | - | 0 |
| Dense | 128 | 8320 |
| Dropout (0.4) | - | 0 |
| Dense | 1 | 129 |

## 4 Experimental evaluation

The experiments presented below were performed on a commodity machine with 12 logical cores (Intel(R) Core(TM) i5-10500H CPU @ 2.5GHz) and 16 GB of RAM, running Windows 11.

### 4.1 Dataset description

The dataset that was used is a collection of the cardiorespiratory measurements acquired during 992 treadmill maximal Graded Exercise Tests (GET) performed in the Exercise Physiology and Human Performance Lab of the University of Malaga. HR, *V O*_2_, carbon dioxide generation, and pulmonary ventilation were measured on a breath-to-breath basis along with the treadmill speed during maximal effort tests. Participants are amateur and professional athletes of ages ranging from 10 to 63 years old. The age, height, and weight of the participants are provided, as well as the temperature and humidity during the test. The dataset can be found here^2^.

### 4.2 Experimental results

To evaluate our proposed NN architecture, we first needed to split the dataset into train and test sets. To this end, we split each subject’s data into 80% for training and 20% for testing. Then, a NN model for each subject was created based on its respective train set and tested against its respective test set. To estimate the NN’s ability to predict *V O*_2_,we used the following metrics:

- Mean Absolute Error (MAE) is an average of the absolute errors |*e*_*i*_| = |*y*_*i*_*− x*_*i*_|, where *y*_*i*_ is the predicted*V O*_2_ and *x*_*i*_ is the actual *V O*_2_.
- Mean Squared Error (MSE) is the average squared difference between the predicted values of *V O*_2_ and the actual ones. This measure has the characteristic of penalizing errors due to the squared difference, thus acting as a measure to distinguish approaches with larger errors in their predictions.

To further investigate the proposed NN’s predictions, we decided to cluster the data before training the network. To do so, we clustered the data into groups based on their age, sex, weight and height. Specifically, the age was split into ten-year segments, the weight into ten-kilo segments and the height into ten-centimeter segments. For instance, two men in their twenties that are between 70 to 79 kilos and 170 to 179 centimeters tall will fall into the same group. Then, for each subject in each group we split the data into train (80%) and test (20%) sets and one NN model was created per group. Furthermore, this “clustered neural network” model used as training the train sets of all the users in the same group and was tested on the remaining test sets of the same group. Finally, both the linear correlation approach and the theoretical method were evaluated on the same test sets with the same metrics as the NNs.

For the evaluation of the predictions in the entirety of the dataset, we used a distribution plot of MAE and MSE which illustrates the distribution of each evaluation metric for each method and for each subject. Figures 1 and 2 illustrate the MAEs and MSEs for all subjects respectively. The more dense the values are to the left of the figures, the lesser the prediction errors are. Lower prediction errors indicate increased prediction performance. From these two figures, we can observe that the neural network approaches outperform both the linear correlation approach and the theoretical method, having the lowest prediction errors. Furthermore, it can be seen that although both neural network approaches pose similar prediction performances, the clustered one performs slightly worse. This can be explained by the fact that the clustered neural network averages out its training based on multiple subjects of its group, therefore it is not fine-tuned for a specific subject but for a specific group. The similarity of their prediction performance though, indicates that a clustered neural network could be used in cases where data for a specific subject are missing.

**Figure 1:**
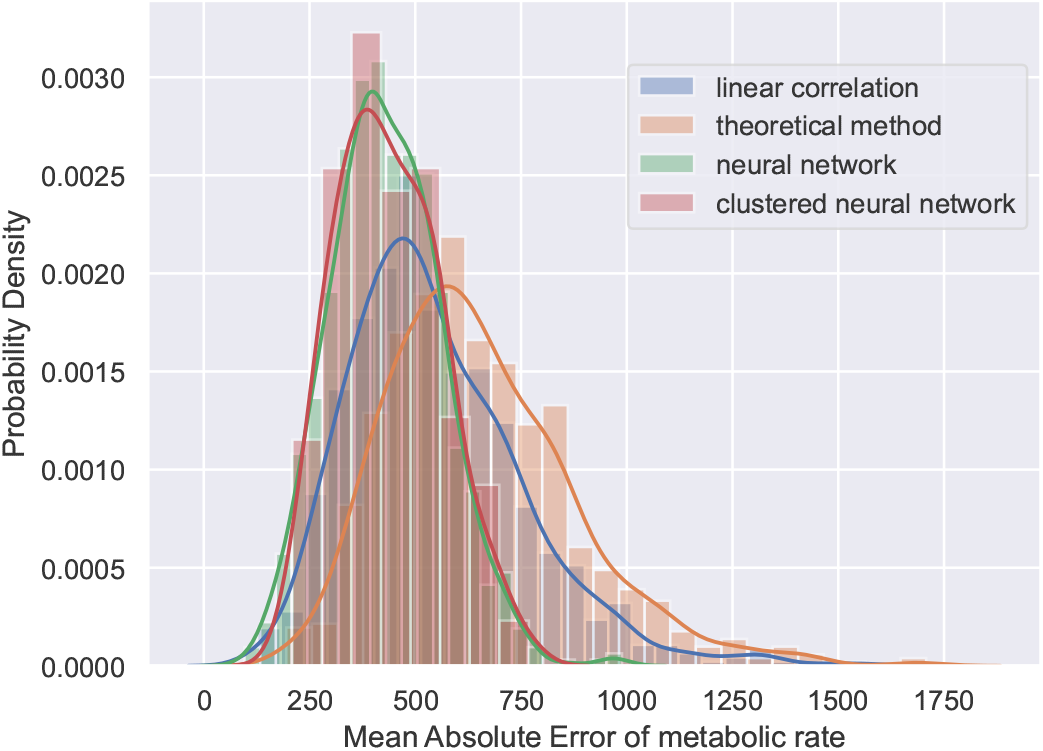
The Mean Absolute Error (MAE) of the methodologies.

**Figure 2:**
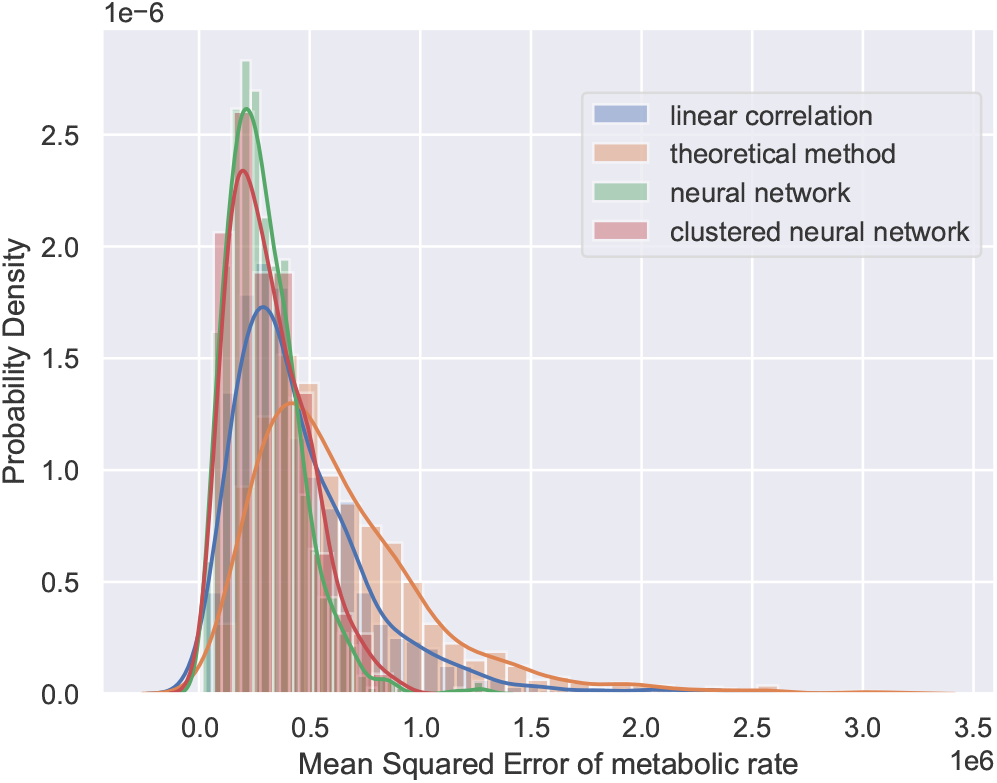
The Mean Squared Error (MSE) of the methodologies.

The last step of the experiments was to compare the execution performance of the NN against the other methodologies and evaluate whether it is able to be deployed in smartwatches, smartbands or smartphones. To do so, we measured mthe time taken to predict the *V* 0_2_ for every methodology; i.e., the wall clock time from the moment the data of the HR and the speed are received to the moment the methodology outputs a prediction. Table 2 presents the findings in milliseconds. We can observe that all of the methodologies take a trivial amount of time to actually predict the *V O*_2_.

**Table 2:** The average execution performance of each methodology in milliseconds.

| NNs training | NNs | Linear correlation | Theoretical method |
| --- | --- | --- | --- |
| 3,330 | 0.14 | 0.0003 | 0.0005 |

We can also observe that the NNs have the worst performance. Nonetheless, the amount of time taken to predict the *V* 0_2_ is only 0.14 milliseconds which indicates that such a prediction model could be used by any wearable device. Finally, the major drawback of the NNs is the training time which takes approximately 3.3 seconds. Be that as it may, such an amount of time can be considered trivial and the NN can be trained offline and given to the user at a later time for real-time predictions.

## 5 Conclusions

In this paper, a NN architecture was proposed that is able to predict a subject’s *V O*_2_. Furthermore, the proposed approach was evaluated on a real-world dataset against two other methodologies based on their ability to accurately predict *V O*_2_. The evaluation used two metrics that estimate the error in the prediction, namely MAE and MSE. The experimental results demonstrated that the proposed NN was able to outperform the other two methodologies. Furthermore, experimental results highlighted the ability of the proposed NN to predict a subject’s *V O*_2_ even when there are no data for the subject but falls into a broader group based on its age, sex, height and weight; factors that can be easily obtained and contribute to a subject’s oxygen uptake. Moreover, execution performance experiments demonstrated that although the neural networks are outperformed by the other methodologies, the amount of time taken to predict the metabolic rate is trivial. This indicates that the NNs can be deployed in wearable devices. Finally, this study provides researchers in the field of nutrition and dietetics insight and an alternate way of measuring a subject’s *V* 0_2_, and subsequently, its metabolic rate.

## Data Availability

All data produced are available online at
https://physionet.org/content/treadmill-exercise-cardioresp/1.0.0/

## Footnotes

1 https://www.iso.org/standard/34251.html

2 https://physionet.org/content/treadmill-exercise-cardioresp/1.0.0/

## References

Sandra Bot and Peter Hollander. The relationship between heart rate and oxygen uptake during non-steady state exercise. Ergonomics, 43:1578–92, 11 2000. doi:10.1080/001401300750004005.

Peter Schantz, Jane Salier Eriksson, and Hans Rosdahl. The heart rate method for estimating oxygen uptake: analyses of reproducibility using a range of heart rates from commuter walking. European Journal of Applied Physiology, 119 (11-12):2655–2671, 2019. ISSN 14396327. doi:10.1007/s00421-019-04236-0.

Scott E. Crouter, Carolyn Albright, and David R. Bassett. Accuracy of polar s410 heart rate monitor to estimate energy cost of exercise. Medicine and; Science in Sports and; Exercise, 36(8):1433–1439, 2004. doi:10.1249/01.mss.0000135794.01507.48.

Kasie D. Cooper and Alex B. Shafer. Validity and reliability of the polar a300’s fitness test feature to predict vo2max. International journal of exercise science, 12(4):393–401, 2019.

Ali Erdogan, Cem Cetin, Hilmi Karatosun, and Metin Lutfi Baydar. Accuracy of the polar s810itm heart rate monitor and the sensewear pro armbandtm to estimate energy expenditure of indoor rowing exercise in overweight and obese individuals. Journal of Sports Science and Medicine, 9(3):508–516, 2010.

Lilian Roos, Wolfgang Taube, Nadja Beeler, and Thomas Wyss. Validity of sports watches when estimating energy expenditure during running. BMC Sports Science, Medicine and Rehabilitation, 9(1), 2017. doi:10.1186/s13102-017-0089-6.

Oludare Isaac Abiodun, Aman Jantan, Abiodun Esther Omolara, Kemi Victoria Dada, Nachaat AbdElatif Mohamed, and Humaira Arshad. State-of-the-art in artificial neural network applications: A survey. Heliyon, 4(11):e00938, 2018. ISSN 2405-8440. doi:https://doi.org/10.1016/j.heliyon.2018.e00938. URL https://www.sciencedirect.com/science/article/pii/S2405844018332067.

Bimal K. Bose. Neural network applications in power electronics and motor drives—an introduction and perspective. IEEE Transactions on Industrial Electronics, 54(1):14–33, 2007. doi:10.1109/TIE.2006.888683.

Ioannis Kontopoulos, Antonios Makris, and Konstantinos Tserpes. A deep learning streaming methodology for trajectory classification. ISPRS Int. J. Geo Inf., 10(4):250, 2021. doi:10.3390/ijgi10040250. URL https://doi.org/10.3390/ijgi10040250.

Antonios Makris, Ioannis Kontopoulos, and Konstantinos Tserpes. COVID-19 detection from chest x-ray images using deep learning and convolutional neural networks. In Constantine D. Spyropoulos, Iraklis Varlamis, Ion Androutsopoulos, and Prodromos Malakasiotis, editors, SETN 2020: 11th Hellenic Conference on Artificial Intelligence, Athens, Greece, September 2-4, 2020, pages 60–66. ACM, 2020. doi:10.1145/3411408.3411416. URL https://doi.org/10.1145/3411408.3411416.

Chiara Zecchin, Andrea Facchinetti, Giovanni Sparacino, Giuseppe De Nicolao, and Claudio Cobelli. Neural network incorporating meal information improves accuracy of short-time prediction of glucose concentration. IEEE Transactions on Biomedical Engineering, 59(6):1550–1560, 2012. doi:10.1109/TBME.2012.2188893.

D. Thangamani and P. Sudha. Identification of malnutrition with use of supervised datamining techniques – decision trees and artificial neural networks. International Journal of Engineering and Computer Science, 3(09), Sep. 2014. URL http://www.ijecs.in/index.php/ijecs/article/view/1546.

HooSeung Na, Joon-Ho Choi, HoSeong Kim, and Taeyeon Kim. Development of a human metabolic rate prediction model based on the use of kinect-camera generated visual data-driven approaches. Building and Environment, 160:106216, 2019. ISSN 0360-1323. doi:https://doi.org/10.1016/j.buildenv.2019.106216. URL https://www.sciencedirect.com/science/article/pii/S0360132319304263.

HooSeung Na and Taeyeon Kim. Development of metabolic rate prediction model using deep learning via kinect camera in an indoor environment. IOP Conference Series: Materials Science and Engineering, 609(4):042036, sep 2019. doi:10.1088/1757-899x/609/4/042036. URL https://doi.org/10.1088/1757-899x/609/4/042036.

Jinsong Liu, Isak Worre Foged, and Thomas B. Moeslund. Automatic estimation of clothing insulation rate and metabolic rate for dynamic thermal comfort assessment. Pattern Analysis and Applications, 2021. doi:0.1007/s10044-021-00961-5.

Patty S. Freedson, Kate Lyden, Sarah Kozey-Keadle, and John Staudenmayer. Evaluation of artificial neural network algorithms for predicting mets and activity type from accelerometer data: validation on an independent sample. Journal of Applied Physiology, 111(6):1804–1812, 2011. doi:10.1152/japplphysiol.00309.2011.

Megan P. Rothney, Megan Neumann, Ashley Béziat, and Kong Y. Chen. An artificial neural network model of energy expenditure using nonintegrated acceleration signals. Journal of Applied Physiology, 103(4):1419–1427, 2007. doi:10.1152/japplphysiol.00429.2007.

P. J. Butler, J. A. Green, I. L. Boyd, and J. R. Speakman. Measuring metabolic rate in the field: the pros and cons of the doubly labelled water and heart rate methods. Functional Ecology, 18(2):168–183, 2004. doi:https://doi.org/10.1111/j.0269-8463.2004.00821.x.

Nils Peter Vilhelm Lundgren. The physiological effects of time schedule work on lumber-workers. Lagerström boktryckare, 1947.

Lr Keytel, Jh Goedecke, Td Noakes, H Hiiloskorpi, R Laukkanen, L Van Der Merwe, and Ev Lambert. Prediction of energy expenditure from heart rate monitoring during submaximal exercise. Journal of Sports Sciences, 23(3): 289–297, 2005. doi:10.1080/02640410470001730089.

Robert Pettitt, Cherie Pettitt, Chad Cabrera, and Steven Murray. A theoretical method of using heart rate to estimate energy expenditure during exercise. International Journal of Sports Science & Coaching - INT J SPORTS SCI COACH, 2, 09 2007. doi:10.1260/174795407782233146.

